# Study of knowledge and practices of local anaesthetic systemic toxicity among Doctors in Sri Lanka

**DOI:** 10.1101/2021.04.19.21255661

**Authors:** B.M. Munasinghe, AG. Arambepola, N. Subramaniam, S. Nimalan, KUIS. Gunathilake, RD. Nissankaarachchi, S K. Karunathilake, BDW. Jayamanne

## Abstract

**Background:** Local anaesthetic systemic toxicity (LAST) could be potentially life threatening. This study focused on describing the knowledge and practices of use of local anaesthetics (LA) among the doctors in Sri Lanka and the ability to detect and manage an event of LAST.

**Materials and methods:** A descriptive cross-sectional study was conducted among doctors in Sri Lanka using an online self-administered questionnaire based on AAGBI guidelines (2010). Descriptive statistics were analyzed by cross-tabulations and presented as numbers and percentages using IBM-SPSS 25.

**Results:** The response rate was 60% out of 600. Majority were males (58%) while 45% of the respondents were anesthetists. Ultrasound was used by 47.4% during LA. The majority considered total body weight for dose calculations. Around 50% of respondents identified bupivacaine as the most cardiotoxic. The majority utilized some form of monitoring and were knowledgeable on identification, prevention and initial management of LAST. Approximately 45% identified Intralipid (ILE) as the definitive treatment of LAST, out of which, 66.8% knew the correct dose and 77.2% and 26.5%, the availability and location of storage, respectively.

**Conclusion:** The basic knowledge on LAST was satisfactory among the respondents. A statistically significant difference on knowledge on maximum safe doses of LA, ILE in established LAST, its dosage and the availability was identified between anaesthetic and non-anaesthetic doctors and post graduate trainees and the rest of the doctors. Overall, significant lapses were noted with regard to the use of total body weight for dose calculations, use of ultrasound during LA administration and dosage, availability and storage of the definitive therapy, ILE, suggesting updates in these key areas.

## Introduction

Local anaesthetic systemic toxicity (LAST) is rare, underdiagnosed and underreported ^[1]^, but could result in serious morbidity and mortality ^[2]^.

Existing literature emphasizes the importance of knowledge on LAST^[3]^. Knowledge and practices in recognizing, preventing and treating LAST is essential in minimizing and ultimately managing an event of LAST.

We reviewed the literature on factors contributing to LAST and management protocols and studied the knowledge and practices among the doctors in our study population regarding identifying, preventing and managing LAST.

## Materials and methods

The study was conducted as a descriptive cross-sectional study among middle and intermediate-grade doctors in Sri Lanka. Considering 20,000 practicing doctors’ population were eligible for our study ^[4]^ and level of awareness on LAST in a regional study among doctors ^[5]^ being around 30% (outcome factor of 30% selected), at 7.5% confidence limit and 95% confidence interval with a design effect (2.0) for cluster sampling, a sample size of 285 was calculated. Following attrition for 20% for non-responders, the minimum sample size required was 342. A self-administered questionnaire(SAQ) was prepared following review of literature and the Association of Anaesthetists of Great Britain and Ireland (AAGBI) guideline on LAST (2010). Face and content validity and appropriateness to culture were assessed and certified by an expert panel. A single-stage cluster sampling method was utilized. Hospitals were chosen randomly. Following establishing remote verified individual communications (via email or social media (WhatsApp/Viber/ Facebook) the questionnaire was distributed. The questionnaire was accessible to the participants only after the consent. The responses were stored in password-protected online cloud. The analysis of data was done with IBM SPSS (version 25) by applying relevant statistical tests accordingly. P < 0.05 was considered as statistically significant.

Ethical approval was obtained from the Ethical review committee of the Sri Lanka Medical Association. (ERC/20/023)

## Results

Out of 600 participants, 360 responded (response rate -60%) where 58.3 % were males (210). Median age was 32 years (Q1=29.7, Q3=34.4, IQR=4.7).

Majority of respondents were experienced as doctors for 2 to 5 years *(Figure 1: Working experience of the respondents as doctors)*.

**Figure 1.**
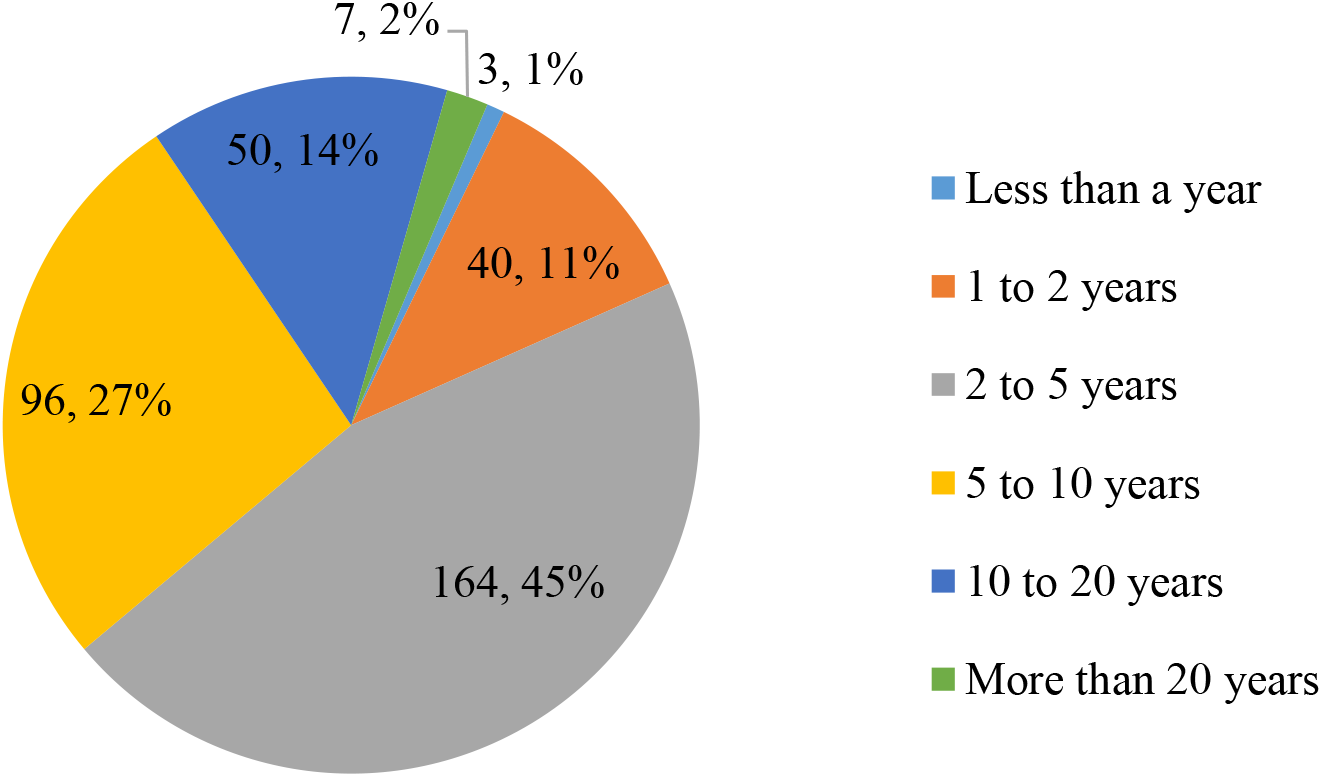
Experience of the respondents as doctors

About 30% were postgraduate trainees from different specialties. (*Figure 2: Categorization according to designation)*.

**Figure 2.**
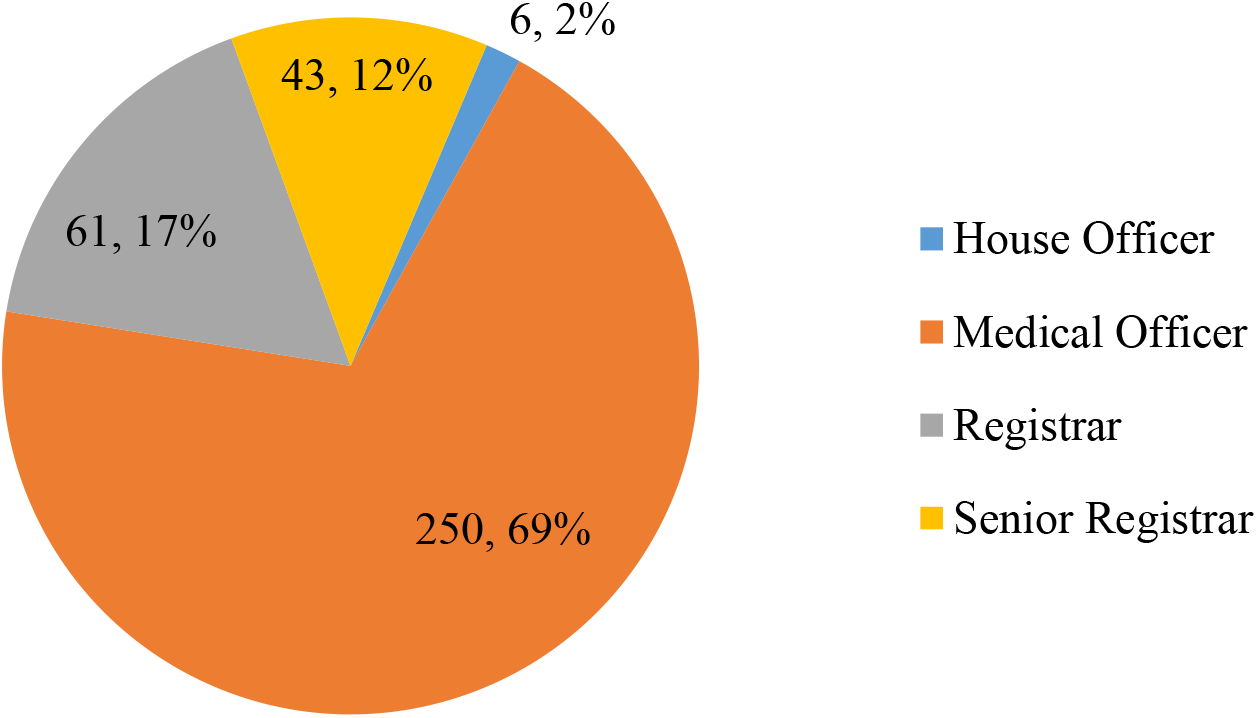
Categorization according to designation

Distribution of responders according to subspecialty is shown in *Table 01*.

**Table 1.** Distribution of responders according to subspecialty

| Subspecialty | Number | % |
| --- | --- | --- |
| Anaesthesia/ ICU | 160 | 44.4 |
| Emergency Medicine | 20 | 5.6 |
| General surgery | 36 | 10.0 |
| Medicine | 39 | 10.8 |
| Gynaecology and Obstetrics | 20 | 5.6 |
| Paediatrics | 12 | 3.3 |
| Ophthalmology | 5 | 1.4 |
| Oral and maxillofacial surgery | 7 | 1.9 |
| Orthopaedics | 6 | 1.7 |
| Radiology | 7 | 1.9 |
| Other* | 48 | 13.4 |
| **Total** | **360** | **100** |

\*ENT, plastic, urology, vascular surgery etc

Distribution of responders according to type of hospital is shown in *Table 2*.

**Table 2.** Distribution of responders according to type of hospital

| Type of hospital | Number | % |
| --- | --- | --- |
| Teaching | 190 | 52.8 |
| Provincial General | 13 | 3.6 |
| District General | 90 | 25.0 |
| Base | 52 | 14.4 |
| Divisional | 8 | 2.2 |
| Other * | 7 | 2.0 |
| **Total** | **360** | **100** |

\*Primary medical care unit/ private dispensary/ private hospital/ preventive sector)

### Practices of use of three local anaesthetic agents (LA)

The frequency and route of usage of the three LA; lignocaine, bupivacaine and prilocaine were studied. Plain Lignocaine was the most commonly used and Prilocaine was the least commonly used. *(Table 03)*.

**Table 3.** Frequency of usage by local anaesthetic agent

| Agent | Frequency of usage per month |  |  |  |  |  |
| --- | --- | --- | --- | --- | --- | --- |
|  | Never(%) | <1(%) | 1-5(%) | 6-15(%) | 16-30(%) | >30(%) |
| **Plain lignocaine** | 25(6.9) | 54(15) | 63(17.5) | 85(23.6) | 61(16.9) | 72(20) |
| **Lignocaine with Adrenaline** | 52(14.4) | 77(21.4) | 88(24.4) | 71(19.7) | 36(10) | 36(10) |
| **Plain Bupivacaine** | 111(30.8) | 36(10) | 65(18.1) | 64(17.8) | 36(10) | 48(13.3) |
| **Prilocaine** | 309(85.8) | 33(9.2) | 09(2.5) | 09(2.5) | 0 | 0 |

### Route

Subcutaneous infiltration was the commonest route (39.7%) followed by regional nerve blocks (39.7%) and epidurals (13%) *(Table 04)*.

**Table 4.** Frequency of usage by route of administration

| Agent | Frequency of usage per month |  |  |  |  |  |
| --- | --- | --- | --- | --- | --- | --- |
|  | Sub-cutaneous infiltration | Regional nerve blocks | Epidural | Intravenous | Retrobulbar/peribulbar | Other (Intercostal/intrapleural) |
| **Plain lignocaine** | 300(43.0) | 185(39.8) | 34(14.9) | 89(80.2) | 35(58.3) | 69(34.5) |
| **Lignocaine with Adrenaline** | 236(33.8) | 107(23.0) | 09(4.0) | 0 | 07(11.7) | 39(19.7) |
| **Plain Bupivacaine** | 154(22.0) | 165(35.5) | 176(77.2) | 07(6.3) | 12(20.0) | 84(42.2) |
| **Prilocaine** | 08( 1.2) | 08( 1.7) | 09(4.0) | 15(13.5) | 06(10.0) | 06(3.0) |
| **Total per month** | **698** | **465** | **228** | **111** | **60** | **192** |
| **Percentage-methods** | 39.7 | 26.4 | 13.0 | 6.3 | 3.4 | 11.2 |

### Usage of ultrasound

Nearly 53% (191) (95% CI 47.7-58.3) never used ultrasound during LA administration *(Figure 3)*. Out of these, 20% (38) were anaesthetists; 16.8% (32), 15.7% (30) and 8.9% (17) were respectively from General Medicine, General Surgery and Gynaecology and Obstetrics, where LA procedures are generally performed in the absence of ultrasound. Around 20% (73) (95% CI 15.78-24.2) responded as using ultrasound ‘always’ or ‘frequently’. Majority (86%, 63) of this category were anaesthetists.

**Figure 3.**
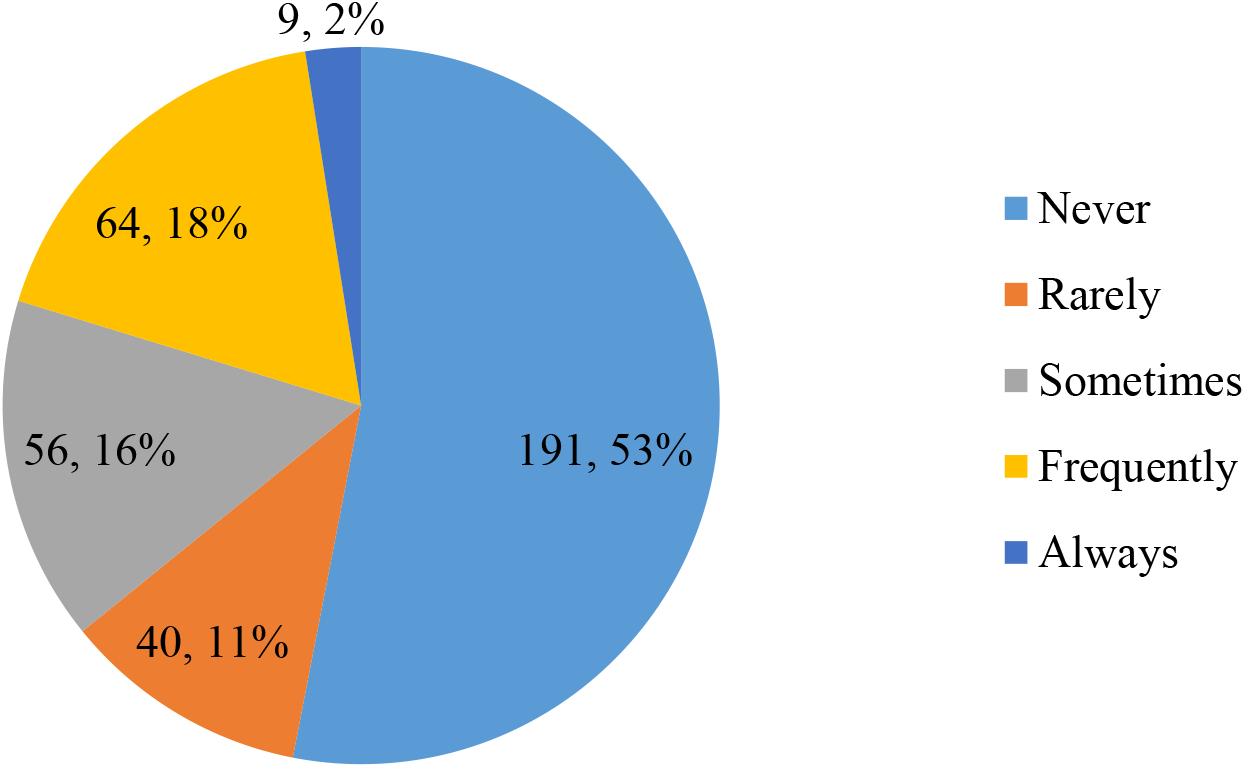
Frequency of usage of ultrasound

With regard to monitoring during LA, the preferred mode was found to be the pulse oximetry. This was utilized by 234 (65%, 95% CI 59.9-70). Out of the participants, 33.3% (120) (95% CI 28.3-38.3), were utilizing all (Pulse oximetry, ECG, Non-invasive blood pressure). Around 14.2% (51) used at least pulse-oximetry while 6.1% (22) opted for non-invasive blood pressure monitoring and 3.3% (12) ECG only. Roughly 23.9% (86) (95% CI 19.4-28.4) did not use any monitoring during LA administration.

A test dose of LA was administered by only around 25% (90) (95% CI 20.4-29.6).

With regard to dose calculations, 42.7% (154) considered the age of the patient and 46.6%(168), the comorbidities while 26.9% (95% CI, 22.2, 31.6) (97) considered ideal or lean body weight. No statistical significance was identified between Anaesthetic(A) vs Non-anaesthetic(NA) (P=0.100), Post-graduate trainees(PG) vs non-PG(NPG) group (P=0.604) and doctors experienced >10 years (E) vs less experienced (NE) (P= 0.835).

The proportions which correctly identified the maximum safe doses of LAs are shown in *Figure 04*.

**Figure 4.**
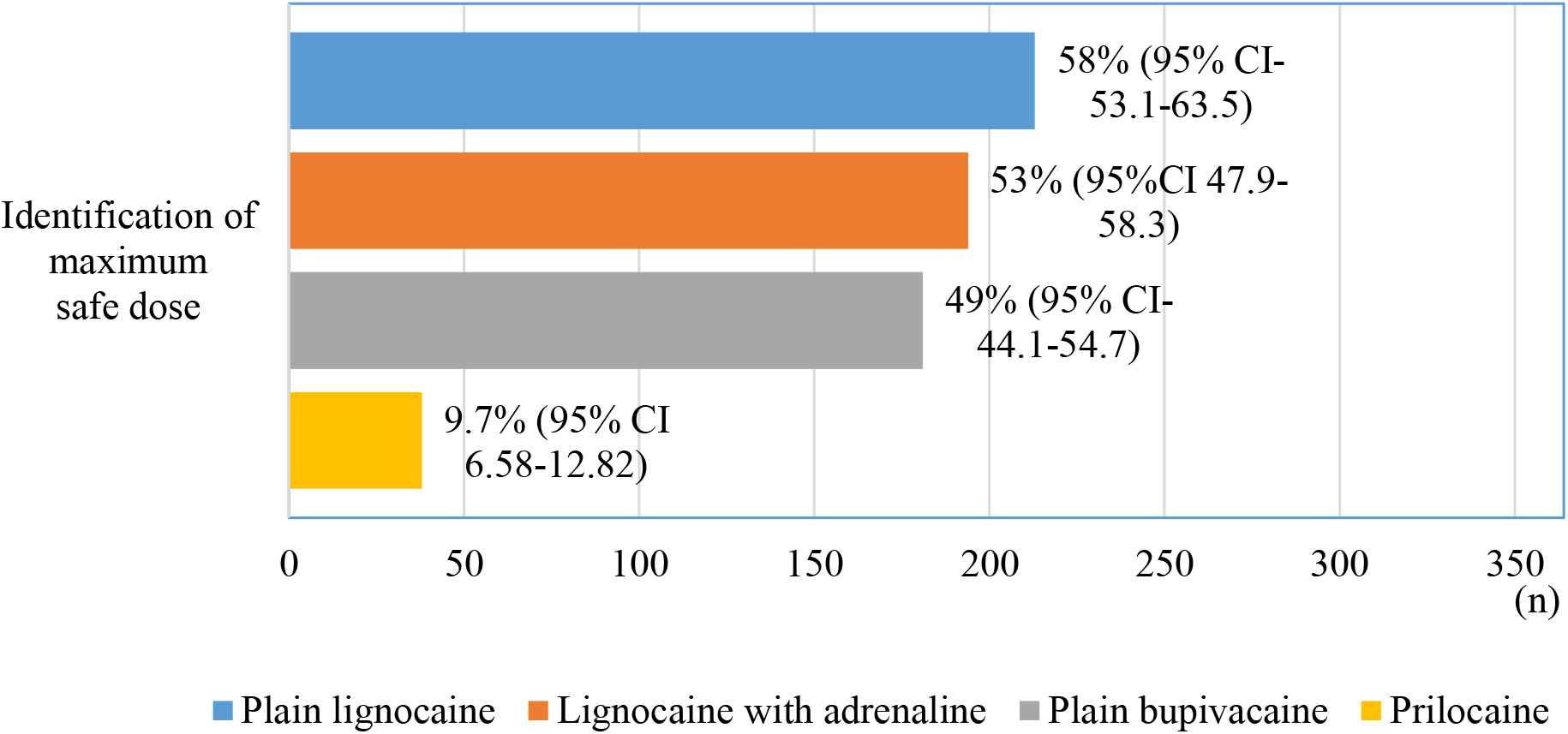
Identification of maximum safe doses of LA

Roughly, 47.8%(172) (95% CI 42.6-53) were able to recognize bupivacaine as the most cardiac toxic out of these. There was a statistically significant difference in the knowledge on cardiotoxicity of bupivacaine between A and NA groups (p<0.001) and E vs NE (P= 0.0003) groups. Knowledge on safe doses was significantly different between PG trainees (PG) and non PG trainees(NG), and Anesthetic (A) and non-anesthetic (NA) doctors (p<0.05) *(Table 5)*.

**Table 5.**
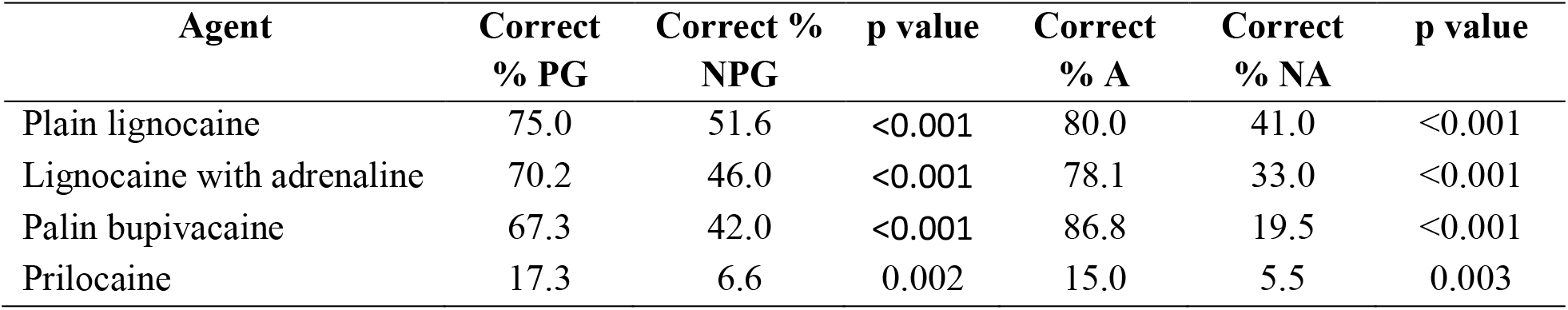
Association of the knowledge of safe doses among PG trainees (PG) and non PG trainees(NG), Anesthetic (A) and non-anesthetic (NA) doctors

Around 93.3% (336) (95% CI 90.6-95.9) had heard about the systemic toxicity of LA and 22.5% (81) (95% CI 18.1-26.9) have witnessed an episode. Approximately 95% (95% CI 92.7-97.3) or more of the participants had knowledge on both cardiovascular (n=352) and neurological features (n= 345) of LAST. Hypertension, (81, 22.5%, 95%CI 18.1-26.9) and Tachycardia (n= 165, 45.8%, 95% CI 40.6-51) were relatively less known clinical features among the respondents. Comparatively higher knowledge was elicited among A group (p=0.05) and PG group (p= 0.01) in comparison to NA and NPG groups respectively regarding these ‘prodromal’ cardiovascular features.

The knowledge on steps which could be effective in minimizing or preventing LAST revealed that 75.3% (271) (95% CI 71-80.2) decided on necessity of monitoring during LA use while 78.33% (282) (95% CI 73.93-82.7) identified aspiration prior to administration of the LA and 65% (234) (95% CI 59.9-70.0) the importance of ultrasound use. Around 42.5% (153) (95% CI 37.3-47.7) accepted that addition of adrenaline to the LA solution could prove to be useful in this aspect. More anaesthetic doctors recognized the former in prevention of LAST with a statistical significance (p= 0.00007) compared to NA group. Similar statistical significance (p=0.0005) was identified between PG and NPG categories.

Approximately, 83% (300) (95% CI 79.0-86.9) respondents identified intravenous fluid and oxygen as part of the management of an established LAST while 57% (205) (95% CI 51.78-62.2) opted for prolonged cardiopulmonary resuscitation. Only 14.7% (53) (95% CI 10.9-18.4) considered cardiopulmonary bypass as part of the therapy. Around 15% (95% CI 11.24-18.7) respondents (55) considered use of intravenous propofol as part of the treatment.

Intravenous ILE as the definitive therapy for LAST was recognized by 45% (162) (95% CI 39.7-50.2) of the respondents. A vs NA and PG vs NPG groups yielded a P value of 0.0001 and E vs NE, this was 0.015 which were significant statistically. Around 66% (107) (95% CI 61.01-70.9) of this group identified the correct dose of the ILE where knowledge of A group was statistically significant over NA group (P=0.001). Approximately 54 (33.3%, 95% CI 28.3-38.3) of this subcategory, responded that ILE was not available in their institution. The respondents who were knowledgeable on the availability (PG vs NPG, P=0.012) (108, 66.6%, 95% CI 61.6-71.6) of the subcategory), 43 (26.5%, 95% CI 21.8-31.1)) knew where it was stored in comparison to 65 (40.1%, 95% CI 34.4-45.8) who did not know the location of storage. A statistical significance (p=0.012) was observed between PG vs NPG subcategories.

## Discussion

Local anaesthetic agents are used frequently in most subspecialties of medicine ^[6]^. The growing interest in regional nerve blocks, enhanced recovery after surgery (ERAS) protocols and multimodal analgesic regimens, have led to an increase in the usage of LA^[7]^. Global increase in aging population may further increase the potential of LAST in the future ^[8]^.

The incidence of LAST is about 0.87 per 1000 peripheral nerve blocks ^[9]^ Epidural blocks are commonly associated with LAST, followed by axillary and interscalene blocks ^[10]^. Most cases reported are related to bupivacaine, attributed to its increased cardiotoxicity. Serious LAST can be as common as epidural haematomas and peripheral nerve injury ^[11]^. LAST could occur after continuous infusions as well as single injections of LA ^[1]^

Interestingly, a case series revealed that around 20% cases of LAST has been occurring outside the conventional hospital setting where 50% involved were non-anaesthetists and 20% of events were following simple infiltration^[11]^.

About 60% of LAST presents with a typical picture comprising of central neurological (CNS) and cardiovascular (CVS) symptoms. Seizures, agitation and loss of consciousness are common CNS symptoms while dysarthria, perioral numbness, confusion, obtundation and dizziness are rare. CNS symptoms typically precede CVS symptoms ^[10]^ which include both tachy and bradyarrhythmias resulting in reduced cardiac output ^[12]^. This results in a vicious cycle of reduced coronary perfusion which can ultimately result in cardiac arrest ^[8]^. However, 40% of patients can present atypically where symptoms can either delay or CVS symptoms can occur without CNS manifestations ^[11]^.

Our study demonstrated that, out of the basic monitoring of ECG, non-invasive blood pressure and peripheral oxygen saturation, around one-quarter (24%) did not establish any monitoring during LA administration and one-third (33.3%) were opting for all three parameters. Monitoring electrocardiography (ECG), pulse oximetry and blood pressure are important in detecting LAST early ^[3]^. Further, monitoring should be continued for some time for recurrent ^[1]^ and late-onset toxicity, especially in cases of continuous infusions ^[13]^. The pulse oximetry was preferred in this study by many probably due to the ease of use.

The study revealed a higher percentage (25%) of participants stating giving a test dose. In contrast, a recent cross-sectional study among emergency physicians in Turkey ^[14]^, demonstrated a relatively lesser proportion (5%) during their analysis. A different study conducted among the ophthalmologists in Turkey revealed that around 97.1% of the participants were not using a test dose ^[15]^. Some authors suggest the use of a test dose particularly during nerve blockade whenever critical LA volumes are used or for normal volumes in patients with co-morbidities, in view of minimizing LAST ^[16]^.

Safe dose of LA is dependent on multiple factors including the agent, the type of block, age of the patient, body weight, comorbidities and physiological variations such as pregnancy ^[3, 8, 13, 17]^. Though there is no clear evidence of exact safe doses of the different agents, maximum safe doses found in literature ^[18]^ are shown in *Table 6*.

**Table 6.** The different agents and maximum safe doses found in literature

| **Local anaesthetic agent** | **Maximum safe dose (mg/kg)** |
| --- | --- |
| Plain lignocaine | 3 |
| Lignocaine with adrenaline | 7 |
| Plain bupivacaine | 2 |
| Prilocaine | 6 |

Maximum safe doses of LA are inconsistent among the medical community. An overall clinical judgement considering demographic variations and clinical state of the patient are more appropriate than a simple dosage recommendation. Dose calculations based on body weight are variable in literature, where ideal body weight is considered by some^[13]^, while lean body weight is preferred by others^[19]^. Total body weight can overestimate total dose in obese and pregnant patients, therefore ideal or lean body weight is more appropriate^[13]^. Among our respondents, only about 27% were using ideal or lean body weight, with no significant association with experience, PG training and between anaesthetists and non-anaesthetists. The proportion, who considered comorbidities, were also just under 50%. The patients with liver, renal, cardiac and central nervous system diseases are at a higher risk of LAST thus doses should be titrated accordingly ^[11]^.

The respondents of this study were commonly using lignocaine and bupivacaine for subcutaneous infiltration and regional blocks. Prilocaine was the least used LA. Considering the ubiquitous use of rest of LA, a significant lapse of knowledge was noteworthy with regard to maximum safe doses.

The relatively infrequent use and unavailability (specially the parenteral preparations) of prilocaine in Sri Lanka probably explain the significantly less (around 10%) knowledge on its safe dose. Just under half identified bupivacaine as the most cardiotoxic. Seven respondents (Paediatrics) considered intravenous use of bupivacaine which could be catastrophic and should be avoided at all times.

Airway management, oxygenation, ventilation and control of seizures are essential components of supportive management in established LAST ^[3, 8]^. Intravenous 20% lipid emulsion (ILE) given as a 1.5ml/kg bolus, followed by an infusion of 15ml/kg/hr is used as specific management. The patient is reassessed at 5-minute intervals and bolus dose is repeated up to 3 times and the infusion rate doubled if toxic features are not resolved ^[20]^. ILE cannot be substituted by propofol ^[3]^. ILE and management protocols should be kept readily available in facilities where local anaesthesia is practiced ^[1, 8]^. On steps to minimize LAST, vital parameter monitoring, aspiration before injection and use of ultrasound were considered by most. Addition of adrenaline to LA was the least preferred. The physiological response to adrenaline (increments of heart rate by around 10 beats/min and blood pressure by 10-15mmHg) ^[21]^ could be an important marker of intravascular injection, particularly as there is a false negative rate of around 2%. ^[11]^ Tachycardia and hypertension, generally considered as cardiovascular prodromal signs were chosen by less number of respondents. Given that the typical pattern of toxicity with preceding neurological features may not be seen among 40% and as the patients could present with cardiovascular collapse ^[10]^, the detection of the former could be decisive. With respect to neurological prodromal symptoms, metallic taste, visual or auditory disturbances were considered by relatively less number of respondents (60%). Overall, a comparative lack of knowledge on the prodromal features of LAST was evident.

Safe administration of LA guided by ultrasonography will further prevent or minimize LAST. Even though, the majority in our study considered ultrasound in minimizing LAST, its practical use was far less. This could be explained by the preference, access to ultrasound and routes of LA administered. For instance, nearly 90% who utilized ultrasound ‘Always’ or ‘Frequently’ were anaesthetists, who perform more regional nerve blocks and access to ultrasound is greater. Use of ultrasound during administration of LA is known to reduce LAST ^[1]^, with evidence of 65% reduction compared to nerve stimulation alone ^[9]^.

The basic management of an established LAST was known by a significantly high number of respondents with majority opting for intravenous fluids and Oxygen. The relatively less proportion (60%) recognized prolonged cardiopulmonary resuscitation (when indicated) where only 13.5% recognized cardiopulmonary bypass as a treatment option. Being an advanced therapy and mostly encountered and taught in the academic scope of postgraduate trainees are most probable reasons for this observation.

The most important aspect of LAST, the definitive therapy with the use of ILE was relatively less known. Some respondents chose propofol but it should be remembered that ILE is not to be substituted by propofol due to relatively low lipid content, potential cardiovascular compromise and the need of larger volumes ^[22]^. Moreover, the knowledge on the correct dose, availability and the location of storage followed the same pattern. Numerous surveys assessing ILE therapy are found in the literature. A study by Edwards et al at demonstrated an overall deficit in knowledge on LAST (including ILE therapy) in a Maternity unit in a UK Hospital. However, teaching programmes led to a significant improvement in knowledge ^[23]^. A Danish study in 2011, conducted among anaesthetists, revealed that around 50% knew about lipid therapy but were not aware how to acquire ILE and that it was not available at places where LA was administered ^[24]^. In a cross-sectional study in Turkey, 42% of the Emergency physicians, correctly identified ILE dosage ^[14]^. A prospective study among junior surgeons and anaesthetists in the UK suggested a significantly lack of knowledge on ILE among the former group (7.3% vs 100%) [25].

The AAGBI, American Society of Regional Anaesthesia and Pain Medicine and American Heart Association ^[26]^, all have endorsed the use of ILE during established LAST. In this modern era of mobile applications and widespread availability of the internet, dosages of most of the drugs could be easily acquired and the dose of ILE in particular, is available in flow charts in most tertiary care centers. Nonetheless, LAST being an emergency where minutes count, quick access to ILE will undoubtedly be decisive.

The subgroup analysis in this study revealed significant statistical differences in knowledge on maximum safe doses of LA, cardiotoxicity of bupivacaine, prodromal cardiovascular features of LAST and addition of adrenaline and performance of a test dose between postgraduate trainees vs non-postgraduate doctors, Anesthetic vs non-anaesthetic doctors and experienced (>10 years) vs less experienced doctors. With regard to the ILE therapy, the correct drug, dosage, availability and location of storage was known with significant statistical significance by anaesthetists and post graduate trainees. The familiarity, increased frequency of LA use, ILE mainly stored in operating theatres and continuous medical education on this aspect among these categories could possibly explain this observation.

## Limitations

The response rate was relatively low for the study. Out of the respondents, almost 50% were anaesthetists. The other subspecialties where LA use is common, in higher concentrations and volumes, such as general surgery, Ophthalmology, Gynaecology and Obstetrics, Orthopaedics and dentistry, the response rate was relatively insufficient. Fields where topical LA is used in higher doses (Respiratory medicine for bronchoscopic procedures for instance), the same pattern was noticed. Even though the frequency of use was assessed, the volumes of LA, were not evaluated. The correct answers were provided following the submission of the questionnaire, although, the reduced response rate of the initial study and the prevailing pandemic (COVID-19) prompted the authors not to proceed with reassessment of knowledge which could have been informative.

## Conclusion

The basic knowledge on LAST was satisfactory among the respondents. Significant lapses were identified with regard to use of total body weight for dose calculations, use of ultrasound during LA administration and dosage and importantly availability and storage of the definitive therapy, ILE.

## Data Availability

The data that support the findings of this study are available from the corresponding author, [MBM], upon reasonable request.

## Recommendations

The authors suggest the following:

- Education programmes on LAST to be conducted specially for the non-anaesthetic doctors who frequently use LA.
- A brief presentation on LAST during academic sessions on regional nerve blocks/ Inclusion of a segment on LAST to the curriculum of the non-anaesthetic post-graduate trainees.
- LAST management protocols to be displayed at locations of LA use. (Operation theatres, OPD theatres, Radiology suites, Dental treatment suites, any other place where high volume LA use is practiced.)
- Make sure that ILE is available in places where LAs used/ Daily checklists/ stocks to be displayed
- Popularize appropriate use of ultrasound use during LA use
- Adverse events during LA use to be reported and audited promptly

## Contributions

MBM and SK conceived the concept for the study. MBM, SN, NS, SG and DN did the literature review. MBM, SK, SN, NS and WJ designed the study. MBM, NS and SN developed the proposal and proceeded to ethical approval. MBM, AA, SN, NS collected data. All authors were involved in data analysis, compiling, and reviewing the final manuscript.

## Acknowledgment

The authors would like to forward their gratitude to all the respondents and Dr. M.D.C.J.P. Jayamanne and Dr. K.A. Jayasundara (Consultant Paediatricians, District General hospital, Mannar, Sri Lanka) for their valuable insights. An initial version of this audit which analyzed the anaesthetic doctors’ data were presented as a free paper in the annual academic sessions of College of Anaesthesiologists and Intensivists of Sri Lankan, 2021.

## Funding

Study was self-funded by the authors No external funding was received.

### Consent

The data were collected only after completion and submission of an on-line consent form which included all the necessary instructions.

### Confidentiality

No data which can lead to potential respondent identification were collected during the study.

